# Persistent Racial, Ethnic, and County-Level Disparities in Invasive Cervical Cancer Incidence in Wisconsin, 1998–2022

**DOI:** 10.1101/2025.06.13.25329584

**Authors:** Madelyn Frey, Laurie Lapp, Lena Swander, Laura Jacques

## Abstract

**Background:** Invasive cervical cancer continues to disproportionately affect minoritized and rural populations. We analyzed racial/ethnic and geographic incidence trends in Wisconsin from 1998-2022.

**Methods:** Incidence rates (per 100,000 individuals with a cervix) were compared by race/ethnicity and county using Wisconsin Cancer Reporting System data.

**Results:** Overall rates declined 28% (8.2 to 5.9/100,000), but unevenly across populations. Compared with Non-Hispanic White individuals, Non-Hispanic Black individuals had nearly double the incidence, and Hispanic individuals–despite a steeper decline (40.8%,15.7 to 9.3/100,000) -continued to have higher rates. Milwaukee County–Wisconsin’s most socially vulnerable county–had among the highest incidence rates and slowest decline over time (19.8%).

**Discussion:** The preventability of invasive cervical cancer makes the persistent disparities in Wisconsin a clear signal of structural inequities demanding urgent attention.

## Background

Invasive cervical cancer is largely preventable, with incidence rates declining significantly since the introduction of Pap smear screening in the 1970s, and further reduced with the advent of high-risk human papilloma virus (hrHPV) testing in 2003 and HPV vaccination in 2006 (quadrivalent) and 2014 (nonavalent).^1-4^ Despite these advances, more than 13,000 new invasive cervical cancer cases and 4,000 deaths are projected in the United States (US) in 2025.^1^ While overall incidence rates have declined since the early 2000’s, rates have plateaued in the last decade and declines have not been equitably distributed. Nationally, vulnerable populations including racially minoritized groups, socioeconomically disadvantaged individuals, uninsured and underinsured individuals, and rural-dwelling individuals bear a disproportionate burden.^5–7^

It is unknown whether similar disparities exist in Wisconsin, a state with some of the most disparate healthcare outcomes in the nation^8^. Our study examines invasive cervical cancer trends in Wisconsin from 1998 (five years prior to the FDA approval of hrHPV testing) to 2022, focusing on differences by race, ethnicity and county.

## Methods

We obtained invasive cervical cancer data from the Wisconsin Cancer Reporting System (WCRS) and national data from the National Cancer Institute (NCI) SEER Incidence Database.^9^ We conducted analyses using SEER*Stat Version 9.0.32.0. We calculated incidence rates using SEER Site Recode for invasive cervical cancer, expressed as observed cases per 100,000 individuals with a cervix and age-adjusted to the 2000 US standard population (Census P25-1130) over 5-year intervals. For inclusivity and conciseness, we use the term “individuals” to refer to people with a cervix, acknowledging that not all individuals at risk for cervical cancer identify as female and we refer to invasive cervical cancer as “cervical cancer” throughout this manuscript. We stratified incidence by race/ethnicity and county at diagnosis. Rates with fewer than 16 cases were suppressed to discourage misinterpretation of unstable data and protect patient confidentiality. We imported the WCRS data into Stata version 18 to generate descriptive tables and figures. WCRS data are categorized by sex assigned at birth.

## Results

### Overall

The overall burden of cervical cancer in Wisconsin decreased 28% (8.2 to 5.9/100,000) from 1,138 cases in 1998-2002 to 921 cases in 2018-2022. The sharpest decline occurred between 1998-2002 and 2003-2007 where the rate decreased 22% (8.2 to 6.4/100,000) (Figure 1).

**Figure 1.**
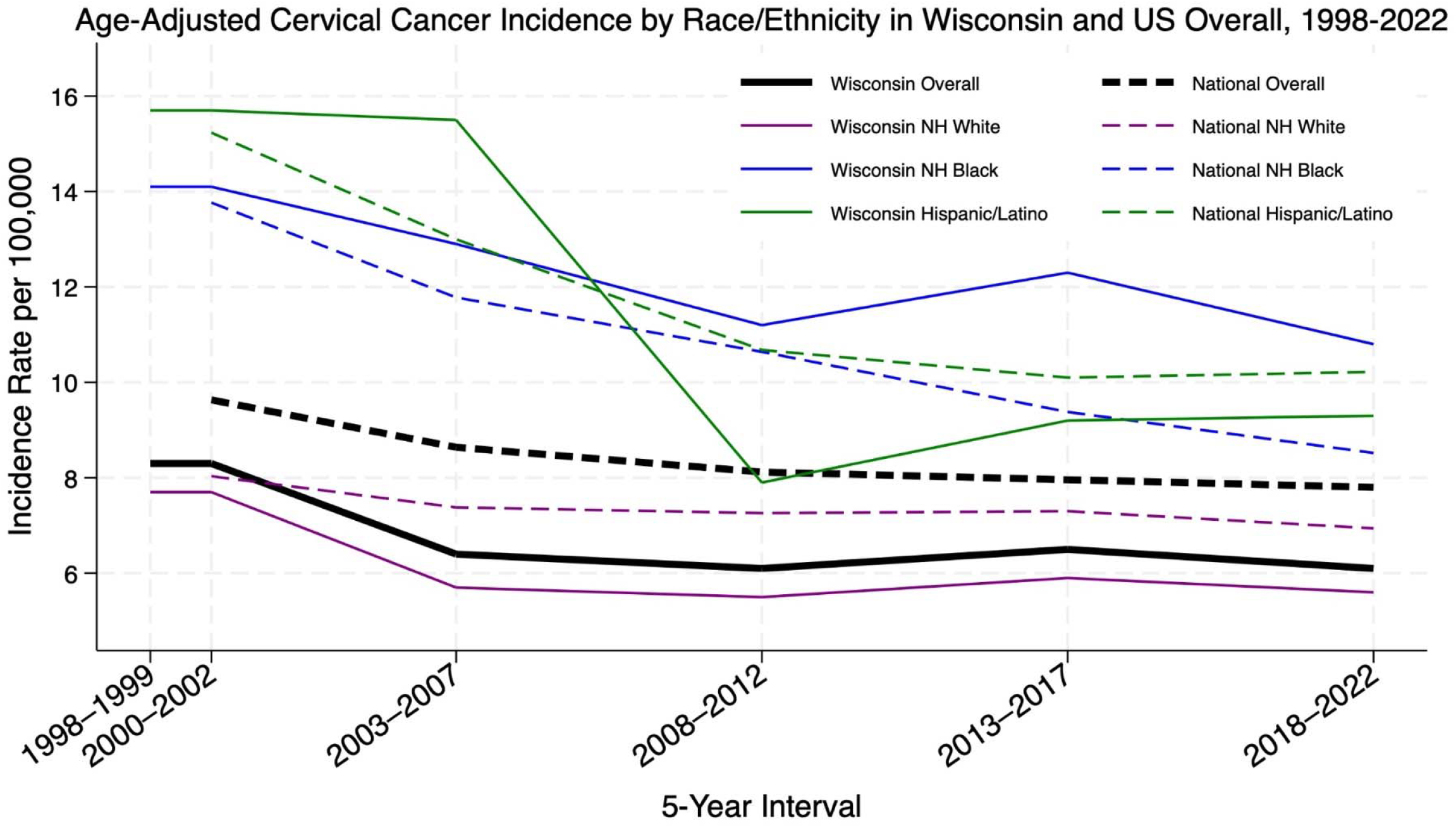
Age-Adjusted Invasive Cervical Cancer Incidence per 100,000 by Race/Ethnicity in Wisconsin and United States, 1998-2022.

### By Race and Ethnicity

Declines in cervical cancer burden in Wisconsin were not evenly distributed across racial and ethnic groups. In Non-Hispanic (NH) White individuals, cervical cancer incidence was the lowest of any measured racial/ethnic group and remained stable throughout the study period. Rates declined from 7.7 to 5.6/100,000 (27.3%) from 1998-2022, and the sharpest decline occurred between 1998-2002 and 2003-2007 (26%; 7.7 to 5.7/100,000). From 2003-2007 onward, rates plateaued (5.6 to 5.9/100,000).

NH Black individuals in Wisconsin had higher cervical cancer incidence rates than the statewide average and nearly twice the rate of NH White individuals for every 5-year interval from 1998 to 2022. The percent difference between NH Black individuals and NH White individuals in Wisconsin grew from 83.1% higher in 1998-2002 to a peak of 119.6% higher in 2013-2017, with a modest decrease to 92.9% higher in 2018-2022 (Figure 1).

Hispanic individuals experienced the largest relative decline in cervical cancer incidence in Wisconsin, decreasing 40.8% (15.7 to 9.3/100,000) between 1998-2002 and 2018-2022. The sharpest drop occurred between 2003-2007 and 2008-2012, with a 49.0% decline (15.5 to 7.9/100,000). However, incidence recently started to rise, increasing 17.7% from 7.9/100,000 in 2013-2017 to 9.3/100,000 in 2018-2022. Hispanic individuals in Wisconsin also experience a disproportionately high incidence of cervical cancer, with the most rate 39.8% higher than that of NH White individuals (Figure 1).

Cervical cancer incidence data for NH American Indian/Alaska Native (AI/AN) individuals were suppressed due to case counts <16 for 1998-2002 and 2018-2022. For the time intervals in which data was available, NH AI/AN individuals had the highest rates of cervical cancer incidence of any race/ethnicity, with incidence rates of 14.4, 19.9, and 14.4 cases per 100,000 in 2003-2007, 2008-2012, and 2013-2017 respectively (Appendix I).

### By County

Among Wisconsin’s three most populous counties (Milwaukee, Dane and Waukesha), Milwaukee County had the highest cervical cancer incidence rate for every five-year interval from 1998-2022, peaking at 10.1/100,000 in 1998-2002 and declining 19.8% to 8.1/100,000 in 2018-2022. Dane County had the second-highest rate for each interval except 2003-2007, when it had the lowest rate of the 3 counties (4.1/100,000). Incidence rates declined 31.9% from 1998-2002 (7.2/100,000) to 2018-2022 (4.9/100,000). Incidence rates in Waukesha County declined 43.1% (6.5 to 3.7/100,000) over the study time period and were below the statewide average for every interval measured (Figure 2).

**Figure 2.**
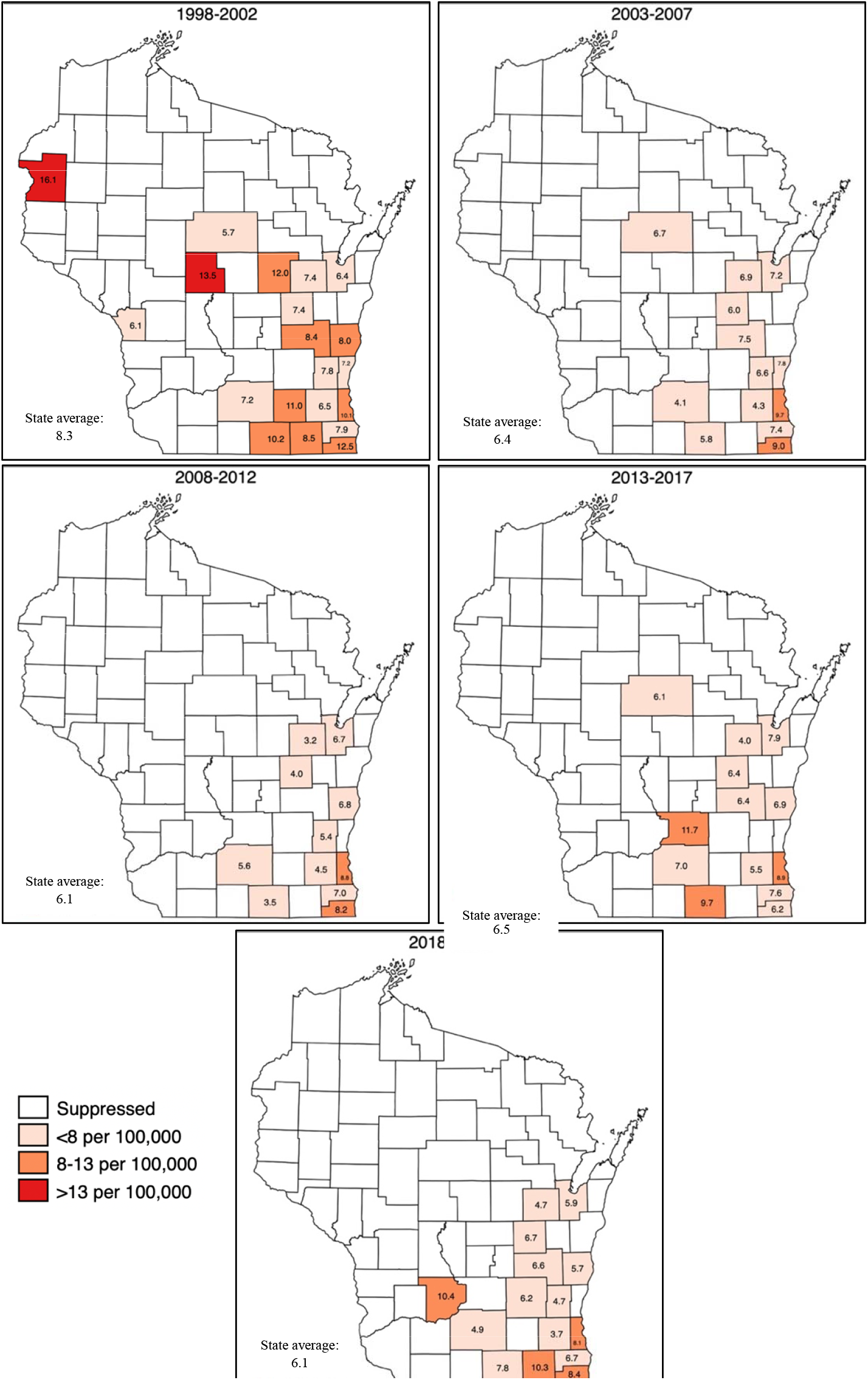
Heat Map of Age-Adjusted Invasive Cervical Cancer Incidence per 100,000 by Wisconsin County, 5-year Intervals (1998-2022)

## Discussion

Cervical cancer is preventable, yet our findings reveal persistent and unacceptable disparities in Wisconsin. While the state has seen an overall decline since 1998, NH Black individuals continue to experience rates nearly double those of NH White individuals, a disparity that has only widened over time. In contrast, the national disparity between NH Black individuals and the NH White individuals narrowed, from 62.8% higher in 1998-2002 to 28.8% higher in 2018-2022 (Figure 1). Notably, this widening disparity in WI has occurred during a period of major public health innovation, including the introduction of hrHPV testing and introduction of the quadrivalent and nonavalent HPV vaccine^1-4^. This inequity cannot be explained by differences in vaccination uptake: state-wide data demonstrates HPV vaccination rates among NH Black adolescents are equal to or higher than those among NH White adolescents.^10^ The growth of the racial/ethnic disparities in Wisconsin, despite comparable vaccination coverage and advances in prevention, contributes to a growing body of evidence that racial disparities in preventable diseases like cervical cancer may not be driven by individual behavior but by systemic racism, the structures, policies and institutions, that systematically advantage certain populations over others.^11,12^

Our data for the three most populous counties in Wisconsin–Milwaukee, Dane, and Waukesha– offer something of a natural case study, given the stark differences in socioeconomic context and structural vulnerability across these settings. The CDC’s Social Vulnerability Index (SVI), which ranks counties from 0 (least vulnerable) to 1 (most vulnerable) based on factors such as socioeconomic and minority status and access to housing and transportation, is a useful measure of the differences.^13^ Dane County ranks near the middle of the state on key structural indicators, including SVI (0.4) poverty rate (10.0%), and median income ($88,108). In contrast, Milwaukee County consistently ranks as the most vulnerable in the state (SVI 1.0), has the highest number of people living below the poverty rate (17.1%), nearly double the statewide average (10.1%), and ranks among the bottom of Wisconsin counties for median household income ($62,118).^13,14^ On the opposite end of the spectrum, Waukesha County has maintained an SVI near 0 across all reporting years, has one of lowest poverty rates (5%) and holds among the highest median household incomes ($104,100).^13,14^

It is therefore notable that Milwaukee County, the most structurally disadvantaged county in the state, also had the highest cervical cancer incidence in every measured time frame and the slowest rate of decline. Furthermore, it cannot be ignored that Waukesha County, one of the most resourced, reliably reports the lowest incidence and the most rapid rate of decline among the three most populous counties. This trend underscores how structural vulnerability and socioeconomic conditions shape disease burden and warrants further research to better understand the drivers of these outcomes and inform the development of effective mitigation strategies.

### Strengths and Weakness

The strengths of our study include the use of long-term, statewide cancer registry data and stratification by race, ethnicity and geography, allowing for a more nuanced understanding of cervical cancer trends. However, several limitations must be acknowledged. Data categorization methods do not fully capture multiracial or ethnically diverse individuals, masking additional disparities and the use of a gender binary in the data categorization methods may also mask additional disparities in certain gender and sexual minorities. Counties with fewer than 16 cases had suppressed rates, which made it difficult to assess trends in the most rural counties. Additionally, we used simple comparisons to describe changes in cervical cancer incidence across five-year intervals over time. While this approach is straightforward and interpretable, it does not allow for statistical testing of changes in trend direction or rate and may overlook inflection points that more advanced methods are designed to detect. Finally, because WCRS data through 2022 are directly observed and not delay-adjusted, there is potential for underreporting in recent years due to reporting lag. This is especially important when comparing to SEER national estimates, which are delay-adjusted, and as such, are not directly comparable.

## Conclusion

Innovation in cervical cancer prevention is accelerating. In-clinic and home-based hrHPV self-collection, extended genotyping, and dual stain cytology will position Wisconsin to meet the World Health Organization’s goal to eliminate invasive cervical cancer by 2030.^15^ But this rapid progress brings urgency: if we fail to address Wisconsin’s persistently wide racial and ethnic disparities now, these advances may not close the gap but widen it.

## Data Availability

All data produced in the present study are available upon reasonable request to the authors

---

**Appendix I. Table 1.**
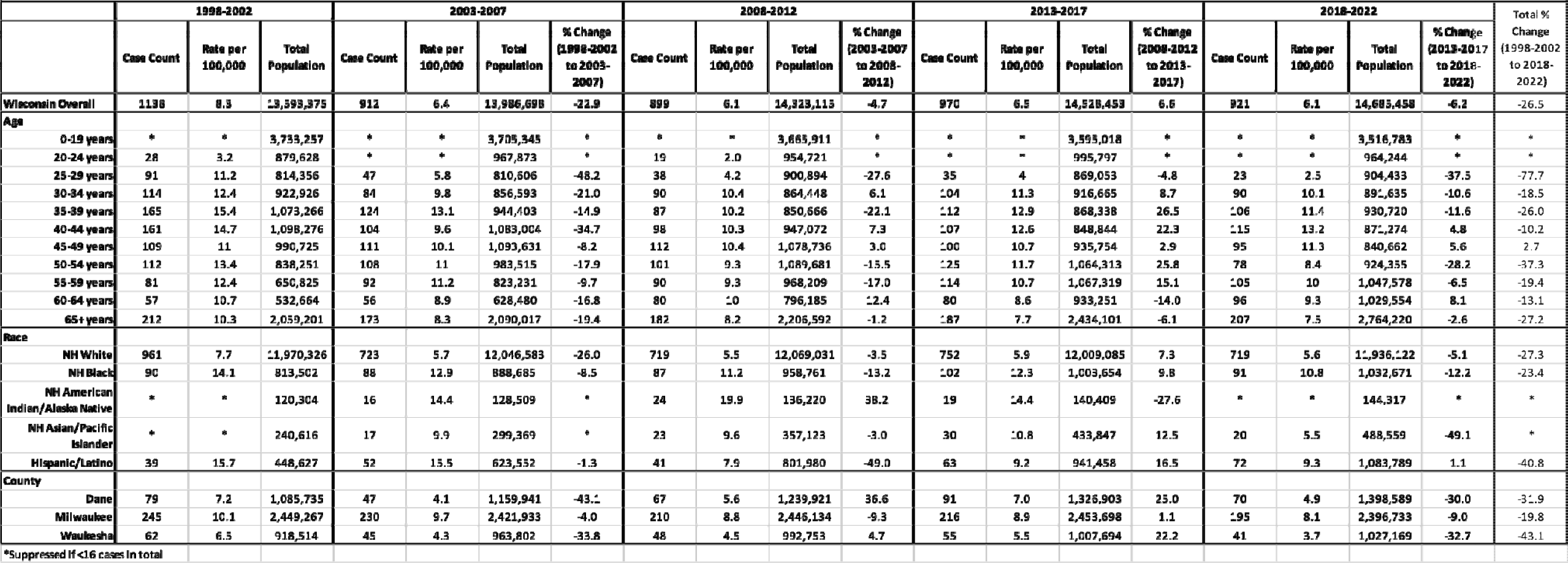
Age-Adjusted Invasive Cervical Cancer Cases, Rates per 100,000 and Percent Change by Age, Race/Ethnicity and Region and Wisconsin Overall, 5-year Intervals (1998-2022)

